# Ongoing HIV transmission following a large outbreak among people who inject drugs in Athens, Greece (2014-2020)

**DOI:** 10.1101/2021.06.24.21258830

**Authors:** Sotirios Roussos, Dimitrios Paraskevis, Mina Psichogiou, Evangelia Georgia Kostaki, Eleni Flountzi, Theodoros Angelopoulos, Savvas Chaikalis, Martha Papadopoulou, Ioanna D Pavlopoulou, Meni Malliori, Eleni Hatzitheodorou, Magdalini Pylli, Chrissa Tsiara, Dimitra Paraskeva, Apostolos Beloukas, George Kalamitsis, Angelos Hatzakis, Vana Sypsa

## Abstract

**Background and Aims:** The HIV outbreak among People Who Inject Drugs (PWID) in Athens, Greece in 2011-2013 was the largest recent epidemic in Europe and North America. We aim to assess trends in HIV prevalence, drug use and access to prevention among PWID in Athens, to estimate HIV incidence and identify risk factors and to explore HIV-1 dispersal using molecular methods during 2014-2020.

**Methods:** Two community-based HIV/hepatitis C programs on PWID were implemented in 2012-2013 (N=3,320) and 2018-2020 (N=1,635) through consecutive Respondent-Driven Sampling (RDS) rounds. PWID were uniquely identified across rounds/programs. We obtained RDS-weighted HIV prevalence estimates per round for 2018-2020 and compared them to 2012-2013. We assessed changes in HIV status, behaviours, and access to prevention in PWID participating in both periods. We estimated HIV incidence in a cohort of seronegative PWID as the number of HIV seroconversions/100 persons-years during 2014-2020 and used Cox regression to identify associated risk factors. Molecular sequencing and phylogenetic analysis were performed in HIV seroconverters.

**Results:** HIV prevalence per round ranged between 12.0%-16.2% in 2012-2013 and 10.7%-11.3% in 2018-2020 with overlapping 95% Confidence Intervals (95% CI). Among PWID participating in both programs, HIV prevalence[95% CI] increased from 14.2%[11.7%-17.1%] in 2012-2013 to 22.0%[19.0%-25.3%] in 2018-2020 (p<0.001). There was a deterioration of socioeconomic characteristics such as homelessness (from 16.2%[95% CI: 13.5%-19.2%] to 25.6%[22.3%-29.0%]), a shift in cocaine use (16.6%[13.9%-19.6%] vs. 28.1%[24.7%-31.7%], reduced access to free syringes (51.8%[48.0%-55.7%] vs. 44.5%[40.7%-48.3%]) and a decrease in daily injecting (36.2%[32.6%-39.9%] vs. 28.5%[25.2%-32.1%]). HIV incidence (95% CI) in 2014-2020 was 1.94 (1.50-2.52) new cases/100 person-years and younger age, lower educational level, larger injection network and daily injecting were risk factors. Almost 9% of HIV seroconversions occurred within a newly expanding phylogenetic cluster.

**Conclusions:** The ongoing HIV transmission among PWID in Athens provides empirical evidence that the current levels of prevention and treatment are inadequate to control the epidemic. Re-evaluation of prevention programs is urgently needed.

## INTRODUCTION

People who inject drugs (PWID) constitute a population with a high burden of HIV infection (1). The first outbreaks among PWID were recorded in the 1980s-1990s in Europe and North America (2). In response to these epidemics, harm reduction services, such as needle and syringe (NSP) and opioid substitution treatment (OST) programs, were introduced and expanded. Since the mid-1990s, no epidemics in Europe were recorded, with the exception of HIV outbreaks occurring in countries of the former Soviet Union (2). The situation changed after 2010 when a series of HIV epidemics were documented once again in Europe and North America (3). Community economic problems, homelessness, and changes in drug injection patterns were factors common to many of these outbreaks (3).

The largest of these outbreaks occurred in Athens, Greece between 2011-2013 (3) where HIV prevalence in the population of PWID increased from less than 1% in 2010 to 16.5% in 2013 (4). This outbreak occurred within the context of low coverage harm reduction programs and financial crisis (5, 6). As soon as it was recognized, there were efforts to expand NSP and OST programs. In addition, a community-based intervention using peer-driven chain referral was implemented in 2012-2013 to recruit a large portion of the PWID population rapidly (estimated population coverage: 88%), test them, and link patients to HIV care (ARISTOTLE program) (4, 7). During ARISTOTLE, HIV incidence decreased rapidly by 78% (4). Since then, there was no systematic implementation of high coverage NSP programs and the number of newly diagnosed HIV cases among PWID in Greece declined but never returned to pre-outbreak levels (8, 9). In 2018-2020, a program with a similar design was implemented in Athens, aiming to enroll a high number of PWID and increase diagnosis and treatment for HIV and hepatitis C infection (ARISTOTLE HCV-HIV).

In this analysis, we combine the data from the two programs in order to: a) assess the change in HIV prevalence, drug use behaviours and access to prevention services from 2012-2013 to 2018-2020), b) estimate HIV incidence and associated risk factors among PWID in Athens during 2014-2020 (i.e., the period following the outbreak), and c) assess the HIV-1 dispersal patterns among PWID and investigate if transmissions continue to occur within the PWID clusters detected during the 2011 outbreak in Athens.

## METHODS

### Participants

Eligible participants of ARISTOTLE and ARISTOTLE HCV-HIV programs were people 18 years or older who had injected drugs in the past 12 months and resided in Athens metropolitan area.

### Design of ARISTOTLE and ARISTOTLE HCV-HIV programs

ARISTOTLE (August 2012-December 2013) and ARISTOTLE HCV-HIV (April 2018-February 2020) were community-based programs aiming at increasing the diagnosis and linkage to care for HIV and HIV/HCV, respectively, in the population of PWID in Athens. A similar design was used in the two programs. Respondent-Driven Sampling (RDS) was used to recruit participants. RDS is a peer-driven chain referral where recruitment begins with a limited number of initial recruits (‘seeds’); individuals receive paper coupons and are asked to draw from their existing social networks to identify up to 3-5 potential recruits, who then present themselves to the program site (10). Coupons include unique identification numbers that allow to link the participant with his or her recruiters and recruits. A dual monetary incentive system was used, in which participants received incentives for participating (primary incentives) as well as for recruiting others (secondary incentives).

Both programs were implemented in multiple consecutive recruitment rounds with a short break in-between; 5 rounds in ARISTOTLE 2012-2013 (of duration 10-12 weeks each) and 2 rounds in ARISTOTLE HCV-HIV 2018-2020 (April 2018-February 2019 and August 2019-March 2020). PWID could participate in multiple rounds but only once in each round. Participants provided the two first initials of their name and surname. These initials along with the full birth date and information on gender were used to identify the participants across recruitment rounds and programs. Identifiers were entered in a database along with other information. When new participants were recruited, their identifiers were checked in the database to make sure they had not participated in the same round. Only twins of the same gender had the same identifier, but this occurred rarely in the two programs. Participants could also provide their full names to facilitate linkage to care.

After obtaining written informed consent, computer-assisted personal interviewing was used to collect information on participants’ sociodemographic characteristics, injection, and sexual behaviour as well as on access to HIV testing, treatment, and prevention programs. A blood sample for HIV testing was collected by the program physician/nurse. As PWID could participate in multiple rounds, multiple HIV tests and questionnaires were available over time for the majority of participants (Figure 1).

**Figure 1.**
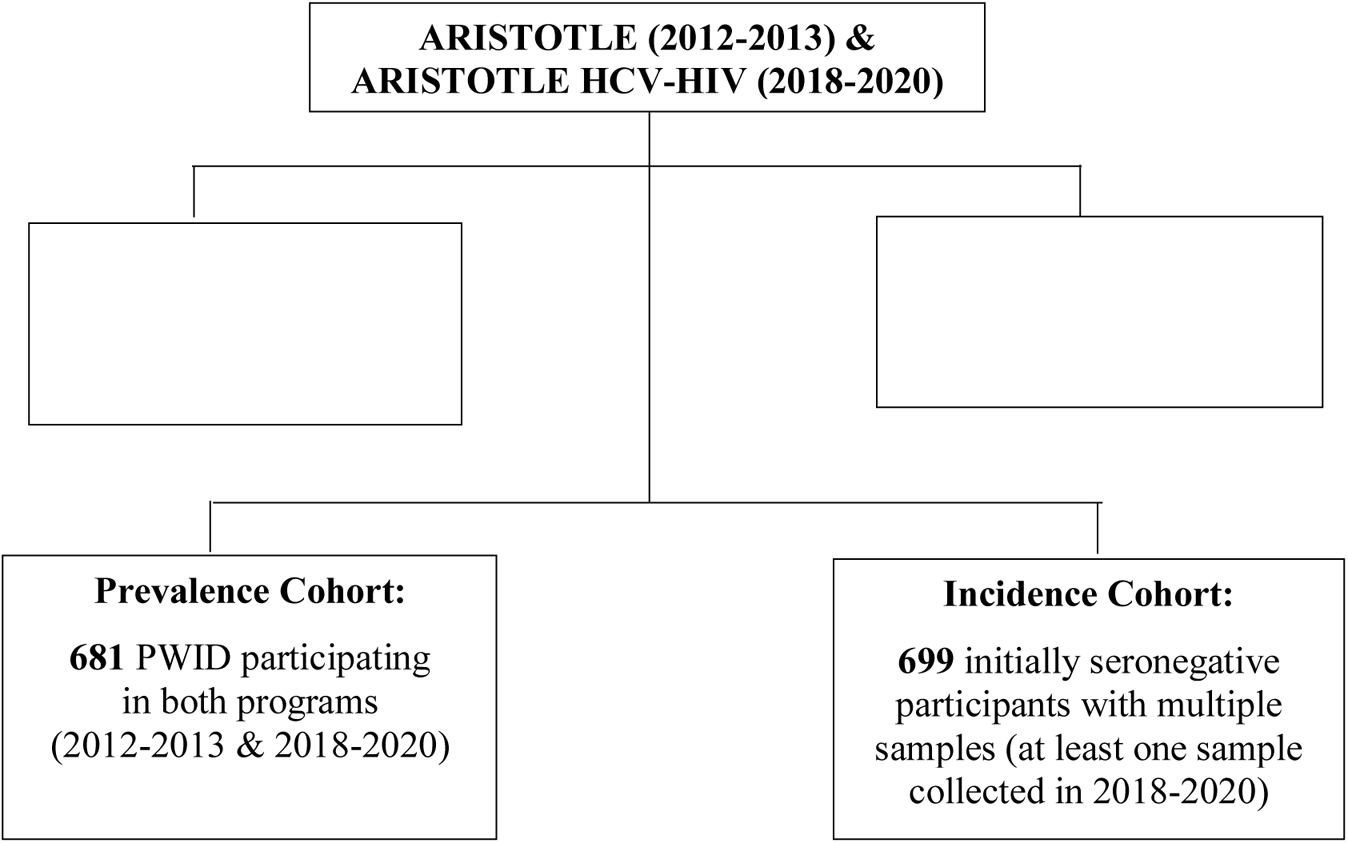
People who inject drugs participating in ARISTOTLE (2012-2013) and ARISTOTLE HCV-HIV (2018-2020) included in the analysis of HIV prevalence and incidence in Athens, Greece.

At the end of this process, participants received coupons to recruit other persons from their injection network as well as their primary monetary incentive. They were asked to return in a few days to collect their secondary incentives and their test result. The program physician/nurse informed the participants about their test results and provided a brief counselling session. HIV cases were reported to the National Public Health Organisation; newly diagnosed cases were identified and prioritised for linkage to HIV care by dedicated staff. In ARISTOTLE HCV-HIV, patients with chronic hepatitis C were linked to HCV care and were followed-up during treatment. Counselling was provided to all participants.

### Outcome measures

The main outcome measure in our analysis was HIV infection status. This was derived from laboratory testing of blood samples using a microparticle enzyme immunoassay anti–HIV-1/2 (AxSYM HIV-1/2 gO; Abbott) and HIV-1/HIV-2 confirmation by Western Blot (MP Diagnostics) or Geenius HIV 1/2 Confirmatory Assay (Biorad).

### HIV prevalence

We obtained HIV prevalence estimates by RDS round in 2018-2020 and compared them to round-specific estimates from ARISTOTLE 2012-2013 (4). RDS Analysis Tool version 7.1 was used to calculate RDS-weighted estimates (11).

In addition, we estimated HIV prevalence in 2012-2013 and 2018-2020 in the subset of PWID who participated in both programs (prevalence cohort, Figure 1) using as numerator the number of PWID who were found to be HIV positive at any participation in each program.

### Trends in socio socio-economic and network characteristics, drug use behaviour and access to HIV prevention services

In the interviews, participants provided information including their injecting network size, sociodemographic characteristics, injecting drug use history, drug treatment, HIV testing experience, and assessment of prevention activities based on a questionnaire adapted from the National HIV Behavioral Surveillance System for PWID in the US (12). We assessed changes in these characteristics over time using data on the first participation of 3,320 and 1,635 PWID in 2012-2013 and 2018-2020, respectively. In addition, we performed this analysis in the prevalence cohort of PWID who had participated in both programs.

### HIV incidence

To estimate HIV incidence during the period 2014-2020, we analysed 699 initially seronegative participants with multiple samples and at least one sample collected in 2018-2020 (incidence cohort, Figure 1). For PWID participating in ARISTOTLE 2012-2013, we considered only those who tested HIV(-) in their last visit to the program as we were interested in seroconversions following the outbreak period. More specifically, we included in the analysis: 1) N=582 PWID for whom the last available HIV test result in 2012-2013 was negative and subsequently participated in one or both rounds in 2018-2020 (2-3 HIV test results available), 2) N=117 PWID who participated only in 2018-2020 (both rounds) and were initially HIV-negative (two HIV test results available) (Table 1).

**Table 1.** Distribution of HIV seroconversions observed during 2014-2020 in a cohort of 699 seronegative community-recruited people who inject drugs in Athens, Greece.

| **ARISTOTLE** | **ARISTOTLE HCV-HIV** |  |  |  |
| --- | --- | --- | --- | --- |
| **2012-2013** | **2018-2020** |  |  |  |
| **Last participation** | **Round A**<br>**April 2018 –**<br>**February 2019** | **Round B**<br>**August 2019 –**<br>**February 2020** | **Number of**<br>**PWID at risk** | **Number of**<br>**Seroconversions** |
| HIV(–) | HIV(+) |  | 343 | 43 |
| HIV(–) |  | HIV(+) | 92 | 3 |
| HIV(–) | HIV(–) | HIV(+) | 147 | 5 |
|  | HIV(–) | HIV(+) | 117 | 6 |
|  | **Total** |  | **699** | **57** |

The HIV incidence rate was calculated as the total number of HIV seroconversions divided by the total person-years at risk (per 100 person-years [PYs]). PWID with a negative HIV test followed by a positive test were defined as HIV-1 seroconverters. The seroconversion time for these subjects was estimated by the midpoint of the interval between the last negative and the first positive test date, as assessed in the two programs. In the process of reporting these new cases to the national HIV surveillance system, we identified a subset of these cases who were already reported with an earlier diagnosis date [30 out of 46 (65.2%) seroconversions that occurred between the two periods]; in this group of PWID, that date was used as the first positive test date. Based on this additional information, we excluded two participants whose estimated seroconversion time was during 2012-2013. For participants who remained HIV negative, person-time at risk was calculated as the interval between the first and the last available sample.

### Sensitivity analysis of HIV incidence estimation

As a sensitivity analysis, we calculated HIV incidence using the random-point method, i.e., assuming seroconversion time to be a random date from a uniform distribution bounded by the last negative and first positive test dates (13–15). To account for the variability of using a random date, multiple iterations were made, and incidence rates were averaged for all iterations.

### Molecular analysis of HIV seroconversions (HIV-1 Subtyping)

Molecular analysis was performed in 35 out of 57 seroconversions in which HIV-RNA was possible to be amplified. HIV-1 subtypes were determined by the automated HIV-1 subtyping tool COMET v0.2 (16) and confirmed by phylogenetic analysis. Specifically, the 35 sequences were analysed phylogenetically along with: i) 247 globally sampled sequences representative of all pure HIV-1 subtypes, sub-subtypes and the majority of circulating recombinant forms (CRFs) collected from the Los Alamos HIV-1 sequence database (http://www.hiv.lanl.gov), and ii) a random collection of 48 sequences from PWID sampled during an outbreak (2011-2015) in Athens, Greece, which fell within the 4 major PWID molecular transmission clusters (MTCs) (subtype A1, subtype B, CRF14_BG, CRF35_AD) (17), used as references. Phylogenetic analysis was performed using the approximate maximum likelihood method (GTR+cat) as implemented in FastTree v2.1 program (18). Furthermore, additional phylogenetic analysis was performed for 3 sequences that did not fall within the 4 PWID MTCs or were not recombinants including partial genomic fragments from the PWID MTCs. Specifically, analysis was performed including a random set of subtype A sequences downloaded from the HV sequence database (N=1,500 sequences) and all available sequences from Greece (N=1,992) sampled since 1999 (17, 19). Phylogenetic tree was reconstructed by using the FastTree v2.1 program as described above. The presence of recombination in all sequences which this procedure did not successfully classify was tested by using the RDP4 (20) and the SimPlot v3.5.1 programs (21). MEGA version X was used to align sequences (MUSCLE algorithm) (22) and FigTree v1.4.3 (http://tree.bio.ed.ac.uk/software/figtree/) to display the annotated phylogenetic trees.

### Statistical analysis

Continuous variables were described using the appropriate measures of central tendency and spread (means and standard deviation (SD) or median, 25^th^ and 75^th^ percentiles). Categorical variables were described using frequencies and percentages. Chi-squared and Mann Whitney U tests or McNemar’s and Wilcoxon signed-rank tests were used to compare the characteristics of participants in the two periods (depending on whether all participants were analysed or only those who had participated in both programs).

Univariable and multivariable Cox proportional hazard models were used to identify factors associated with HIV seroconversion. Hazard ratios and 95% confidence intervals (95% CI) are reported. Participants without seroconversion were censored at the time last known to be event free. Graphical methods as well as tests based on the Schoenfeld residuals (23) were used to check the assumption of proportionality of the hazards. Collett’s approach was applied for model selection and likelihood ratio tests were used for variable inclusion/exclusion decisions (24).

To assess differential loss to follow-up, we compared the characteristics of PWID who participated in both programs versus those who participated only in ARISTOTLE 2012-2013 using t-test, chi-squared test, and Mann-Whitney test, as appropriate.

Analyses were conducted using Stata version 16.0 (25). Two-tailed tests and a significance level of 0.05 were applied; 95% confidence interval (95% CI) are reported. Complete case analysis was used (less than 0.8% of the data was missing in the analysed variables). This analysis was not pre-registered, and results presented in this study should be considered exploratory.

### Ethical Issues

The survey protocols and informed consent forms of the two programs were approved by the Institutional Review Boards of: i) the Medical School of the National and Kapodistrian University of Athens and ii) the Hellenic Scientific Society for the Study of AIDS, STDs, and Emerging Diseases. Eligible persons were asked to provide written informed consent.

## RESULTS

### Participants

Overall, 3,320 and 1,635 PWID were enrolled in ARISTOTLE 2012-2013 and ARISTOTLE HCV-HIV 2018-2020, respectively. In total, 4,274 unique participants were recruited; of those, 681 (15.9%) participated in both programs (prevalence cohort) (Figure 1).

The mean (SD) age of the participants was 35.8 (8.3) and 39.2 (8.3) years in 2012-2013 and 2018-2020, respectively, and the majority were men (84.5% and 83.6%, respectively). They were predominantly active PWID (injection in the past 30 days: 81.2% and 74.9%, respectively) and of Greek origin (83.6% and 84.4%) (Supplementary Table 1). PWID who participated in both programs were more often male, of Greek origin, with stable accommodation, reporting more often history of imprisonment, and sharing syringes as compared to PWID who participated only in 2012-2013 (Supplementary Text, Supplementary Table 2).

### Trends in socioeconomic characteristics, injection practices, access to testing and prevention (2012-2013 and 2018-2020)

The trends in socioeconomic characteristics, injection practices and access to HIV testing and prevention services in PWID who participated in both periods are depicted in Table 2. PWID in 2018-2020 were more frequently homeless (25.6% vs. 16.2%, p<0.001), unemployed (91.0% vs. 78.9%, p<0.001) and without health insurance (79.5% vs. 61.7%, p<0.001), as compared to 2012-2013. There were significant increases in the use of cocaine (28.1% vs. 16.6%, p<0.001) and speedball (14.8% vs. 2.1%, p<0.001) as well as a decrease in the use of heroin (55.3% vs. 80.7%, p<0.001). There was a significant decrease in risky behaviours such as daily injecting drug use (past 12 months: 28.5% vs. 36.2%, p=0.001) whereas syringe sharing remained similar (about half the time or more in the past 12 months: 6.2% vs. 8.5%, p=0.074).

**Table 2.** Trends in socio-economic and network characteristics, drug use behaviour and access to HIV prevention services in people who inject drugs participating in both ARISTOTLE (2012-2013) and ARISTOTLE HCV-HIV (2018-2020) programs (N=681) (as assessed in their first visit to each program).

|  | 2012-2013 | 2018-2020 | p-value |
| --- | --- | --- | --- |
| **A. Socio-economic and network characteristics** |  |  |  |
| Homeless now, n [%; (95% CI)] | 110<br>[16.2 (13.5-19.2)] | 174<br>[25.6 (22.3-29.0)] | <0.001<sup>1</sup> |
| Unemployment, n [%; (95% CI)] | 535<br>[78.9 (75.6-81.9)] | 619<br>[91.0 (88.6-93.1)] | <0.001<sup>1</sup> |
| Without health insurance, n [%; (95% CI)] | 418<br>[61.7 (58.0-65.4)] | 538<br>[79.5 (76.2-82.5)] | <0.001<sup>1</sup> |
| History of imprisonment (past 12 months), n [%; (95% CI)] | 148<br>[21.9 (18.8-25.2)] | 87<br>[12.8 (10.4-15.5)] | <0.001<sup>1</sup> |
| Size of participant's injection network, median (25th, 75th) | 20 (10, 50) | 20 (10, 50) | 0.015<sup>2</sup> |
| **B. Injecting drug use behaviour** |  |  |  |
| Main substance of use (past 12 months), n [%; (95% CI)] |  |  |  |
| Heroin/Thai | 549<br>[80.7 (77.6-83.6)] | 362<br>[55.3 (51.4-59.1)] | <0.001<sup>1</sup> |
| Cocaine | 113<br>[16.6 (13.9-19.6)] | 184<br>[28.1 (24.7-31.7)] | <0.001<sup>1</sup> |
| Speedball | 14<br>[2.1 (1.1-3.4)] | 97<br>[14.8 (12.2-17.8)] | <0.001<sup>1</sup> |
| Other | 4<br>[0.6 (0.2-1.5)] | 12<br>[1.8 (1.0-3.2)] | 0.057<sup>3</sup> |
| Injecting drug use (past 30 days), n [%; (95% CI)] | 554<br>[81.7 (78.6-84.6)] | 536<br>[78.8 (75.6-81.8)] | 0.176<sup>1</sup> |
| Daily injecting drug use (past 12 months), n [%; (95% CI)] | 246<br>[36.2 (32.6-39.9)] | 194<br>[28.5 (25.2-32.1)] | 0.001<sup>1</sup> |
| Receptive syringe-sharing about half the time or more (past 12 months), n [%; (95% CI)] | 58<br>[8.5 (6.6-10.9)] | 42<br>[6.2 (4.5-8.2)] | 0.074<sup>1</sup> |
| Use drugs divided with a syringe that someone else had already injected with about half the time or more (past 12 months), n [%; (95% CI)] | 60<br>[8.9 (6.9-11.3)] | 50<br>[7.4 (5.5-9.6)] | 0.317<sup>1</sup> |
| **C. Access to testing, drug treatment and prevention** |  |  |  |
| Currently in opioid substitution treatment, n [%; (95% CI)] | 93<br>[13.8 (11.3-16.7)] | 207<br>[30.5 (27.1-34.2)] | <0.001<sup>1</sup> |
| Received free syringes (past 12 months), n [%; (95% CI)] | 352<br>[51.8 (48.0-55.7)] | 303<br>[44.5 (40.7-48.3)] | 0.003<sup>1</sup> |
| Tested for HIV (past 12 months), n [%; (95% CI)] | 344<br>[50.9 (47.0-54.7)] | 310<br>[49.4 (45.5-53.4)] | 0.759<sup>1</sup> |

<sup>1</sup>McNemar's test, <sup>2</sup>Wilcoxon signed-rank test, <sup>3</sup>Exact McNemar's test - Abbreviation:
CI, confidence interval.

An increasing percentage of PWID reported being currently in OST (2018-2020 vs. 2012-2013: 30.5% vs. 13.8%, p<0.001); however, there was a decline in the proportion reporting access to free syringes (past 12 months: 44.5% vs. 51.8%, p=0.003) whereas access to HIV testing remained similar (test in the past 12 months: 49.4% vs. 50.9%, p=0.759).

These trends were similar when all participants of the two programs were included in the comparison (Supplementary Table 1).

### Trends in HIV prevalence (2012-2013 and 2018-2020)

In the sample of 681 PWID participating in both periods, HIV prevalence (95% CI) increased from 14.2% (11.7%-17.1%) in 2012-2013 to 22.0% (19.0%-25.3%) in 2018-2020 (p<0.001), i.e., an increase of 54.9% (Table 3). This increase was apparent among both men and women (Table 3).

**Table 3.** Trends in HIV prevalence in people who inject drugs participating in both ARISTOTLE 2012-2013 and ARISTOTLE HCV-HIV 2018-2020 programs in Athens, Greece (N=681).

|  | N | 2012-2013 |  | 2018-2020 |  | p-value<sup>1</sup> |
| --- | --- | --- | --- | --- | --- | --- |
|  |  | HIV(+) | % (95% CI) | HIV(+) | % (95% CI) |  |
| Total | 681 | 97 | 14.2 (11.7-17.1) | 150 | 22.0 (19.0-25.3) | <0.001 |
| Male | 554 | 82 | 14.8 (11.9-18.0) | 126 | 22.7 (19.3-26.5) | <0.001 |
| Female | 127 | 15 | 11.8 (6.8-18.7) | 24 | 18.9 (12.5-26.8) | 0.004 |

<sup>1</sup>McNemar's test - Abbreviation: CI, confidence interval.

When all participants were included in the analysis, HIV prevalence per round ranged between 12.0% and 16.2% in 2012-2013 and 10.7%-11.3% in 2018-2020 with overlapping 95% CIs (Supplementary Table 3).

### HIV incidence (2014-2020)

In the cohort of 699 initially seronegative PWID, there were 57 seroconversions identified during follow up: 46 seroconversions in PWID who tested HIV(-) in 2012-2013 and were found HIV(+) in the first or second round of ARISTOTLE HCV-HIV 2018-2020 (43 and 3, respectively) and 11 seroconversions in PWID who tested HIV(-) in the first round of ARISTOTLE HCV-HIV 2018-2020 and were HIV(+) in the second round (Table 1). Out of 46 PWID testing negative in 2012-2013 and positive in 2018-2020, 30 were already reported to the national HIV surveillance system with an earlier diagnosis date; that date was used as the first positive date.

The overall HIV incidence (95% CI) for the period 2014-2020 was 1.94 (1.50-2.52) new cases per 100 PYs(Table 4).As a sensitivity analysis, HIV incidence (95% CI) was estimated under the random point approach and the results were similar [1.74 (1.33-2.29) new cases/100 PYs].

**Table 4.** HIV incidence and predictors of HIV seroconversion in people who inject drugs, Athens, Greece from a Cox proportional hazards model (N=699 initially seronegative people who inject drugs with multiple samples and at least one sample collected in 2018-2020). The seroconversion date was estimated using the midpoint approach.

| Variable | N (%) | Events | PYs | Incidence/100 PYs (95% CI) | Crude HR (95%CI) | Adjusted HR (95% CI) |
| --- | --- | --- | --- | --- | --- | --- |
| **All participants** | 699 (100) | 57 | 2,932 | 1.94 (1.50,2.52) |  |  |
| **A. Socio-demographic and network characteristics** |  |  |  |  |  |  |
| **Age (years)** |  |  |  |  |  |  |
| <35 | 352 (50.4) | 37 | 1,524 | 2.43 (1.76,3.35) | 1 |  |
| ≥35 | 347 (49.6) | 20 | 1,409 | 1.42 (0.92,2.20) | 0.58 (0.34,1.00)<sup>†</sup> | 0.94 (0.90,0.98)<sup>†</sup> |
| **Gender** |  |  |  |  |  |  |
| Female | 126 (18.0) | 9 | 558 | 1.61 (0.84,3.10) | 1 |  |
| Male | 573 (82.0) | 48 | 2,374 | 2.02 (1.52,2.68) | 1.24 (0.61,2.52) |  |
| **Country of origin** |  |  |  |  |  |  |
| Greece | 653 (93.6) | 54 | 2,740 | 1.97 (1.51,2.57) | 1 |  |
| Other | 45 (6.4) | 3 | 188 | 1.59 (0.51,4.94) | 0.82 (0.26,2.61) |  |
| **Highest level of education completed** |  |  |  |  |  |  |
| High school or higher | 293 (42.2) | 12 | 1,240 | 0.97 (0.55,1.70) | 1 | 1 |
| Middle/secondary school or below | 401 (57.8) | 44 | 1,670 | 2.63 (1.96,3.54) | 2.71 (1.43,5.13) | 2.27 (1.20,4.32) |
| **Homeless<sup>††</sup>** |  |  |  |  |  |  |
| No | 510 (73.1) | 40 | 2,125 | 1.88 (1.38,2.57) | 1 |  |
| Yes | 188 (26.9) | 17 | 803 | 2.12 (1.32,3.40) | 1.12 (0.64,1.98) |  |
| **Currently health insurance** |  |  |  |  |  |  |
| Yes | 256 (36.7) | 18 | 1,132 | 1.59 (1.00,2.52) | 1 |  |
| No | 442 (63.3) | 39 | 1,796 | 2.17 (1.59,2.97) | 1.33 (0.76,2.33) |  |
| **Unemployed** |  |  |  |  |  |  |
| Yes | 586 (84.2) | 44 | 2,442 | 1.80 (1.34,2.42) | 1 |  |
| No | 110 (15.8) | 13 | 477 | 2.73 (1.58,4.70) | 1.51 (0.82,2.81) |  |
| **History of imprisonment<sup>††</sup>** |  |  |  |  |  |  |
| No | 593 (85.1) | 47 | 2,465 | 1.91 (1.43,2.54) | 1 |  |
| Yes | 104 (14.9) | 10 | 459 | 2.18 (1.17,4.05) | 1.14 (0.58,2.26) |  |
| **Size of participant's network** |  |  |  |  |  |  |
| ≤10 | 219 (31.4) | 9 | 922 | 0.98 (0.51,1.88) | 1 | 1 |
| >10 | 478 (68.6) | 48 | 2,004 | 2.40 (1.80,3.18) | 3.13 (1.46,6.71) | 2.21 (1.08,4.51) |

| **B. Injecting drug use behaviour** |  |  |  |  |  |  |
| --- | --- | --- | --- | --- | --- | --- |
| **Main substance of use<sup>††</sup>** |  |  |  |  |  |  |
| Cocaine | 153 (22.0) | 11 | 624 | 1.76 (0.98,3.18) | 1 |  |
| Heroin/Thai | 508 (72.9) | 45 | 2,188 | 2.06 (1.54,2.75) | 1.18 (0.61,2.27) |  |
| Other | 36 (5.2) | 1 | 110 | 0.91 (0.13,6.48) | 0.49 (0.06,3.81) |  |
| **Injecting drug use in the past 30 days** |  |  |  |  |  |  |
| No | 145 (20.9) | 5 | 620 | 0.81 (0.34,1.94) | 1 |  |
| Yes | 550 (79.1) | 52 | 2,292 | 2.27 (1.73,2.98) | 2.77 (1.11,6.93) |  |
| **Duration of injecting drug use (years)** |  |  |  |  |  |  |
| <14 | 356 (51.1) | 37 | 1,520 | 2.43 (1.76,3.36) | 1 |  |
| ≥14 | 340 (48.9) | 20 | 1,402 | 1.43 (0.92,2.21) | 1.32 (0.72,2.41) |  |
| **Daily injecting drug use<sup>††</sup>** |  |  |  |  |  |  |
| No | 522 (74.9) | 29 | 2,251 | 1.29 (0.90,1.85) | 1 | 1 |
| Yes | 175 (25.1) | 28 | 677 | 4.14 (2.86,5.99) | 3.08 (1.83,5.18) | 2.54 (1.50,4.31) |
| **Receptive syringe sharing<sup>††</sup>** |  |  |  |  |  |  |
| Never or rarely | 662 (95.1) | 52 | 2,793 | 1.86 (1.42,2.44) | 1 |  |
| About half the time or more | 34 (4.9) | 5 | 133 | 3.76 (1.56,9.03) | 1.98 (0.79,4.96) |  |
| **Use drugs divided with a syringe that someone else had already injected with<sup>††</sup>** |  |  |  |  |  |  |
| Never or rarely | 665 (95.6) | 51 | 2,826 | 1.80 (1.37,2.37) | 1 |  |
| About half the time or more | 31 (4.5) | 6 | 97 | 6.19 (2.78,13.77) | 3.26 (1.40,7.61) |  |
| **Currently on OST** |  |  |  |  |  |  |
| Yes | 132 (18.9) | 6 | 557 | 1.08 (0.48,2.40) | 1 |  |
| No | 565 (81.1) | 51 | 2,367 | 2.15 (1.64,2.84) | 0.51 (0.22,1.18) |  |

<sup>†</sup>per year, <sup>††</sup>past 12 months - Abbreviations: PYs, person-years; CI, confidence interval; HR, hazard ratio.

### Risk factors for HIV seroconversion

Table 4 presents univariable and multivariable analysis of risk factors for HIV seroconversion (Supplementary Figure 1 for the evaluation of the proportional hazards assumption). Sociodemographic factors including younger age, lower educational level, and larger size of injection network were identified as independent risk factors for HIV seroconversion in the final model. PWID with lower educational level and larger injecting networks had more than two times higher risk of seroconversion. Concerning injection practices, PWID reporting daily injecting in the past 12 months had 2.54 times higher risk of seroconversion as compared to those injecting less frequently (95% CI: 1.50-4.31, p=0.001).

### HIV-1 Subtyping

Subtyping analysis showed that 80% (n=28) of the sequences fell within the previously identified PWID MTCs in Athens (CRF14_BG: n=21, 60%; CRF35_AD: n=4, 11.4%; subtype B: n=2, 5.7%; subtype A: n=1, 2.9%) (Figure 2A). Unique recombinant forms of the virus were also detected (n=4, 11.4%) (Figure 2A). These forms consisted of partial sequences from previously identified PWID MTCs. The remaining sequences (n=3, 8.6%) were classified as sub-subtype A6 and found to belong within a single PWID MTC consisting of sub-subtype A6 sequences from PWID diagnosed during 2014-2019 (Figure 2B). Specifically, this cluster included 10 sequences of which 2 were sampled in 2014 and the remaining 8 were sampled later between 2018 and 2020 (Figure 2B). Within A6 cluster seven sequences were from PWID of Greek ethnicity, and 3 were found to belong to PWID from Albania, Kazakhstan, and Pakistan. One out of these 3 seroconversions classified in this cluster occurred between September 2018 and November 2019 (last negative and first positive test result, respectively).

**Figure 2A.**
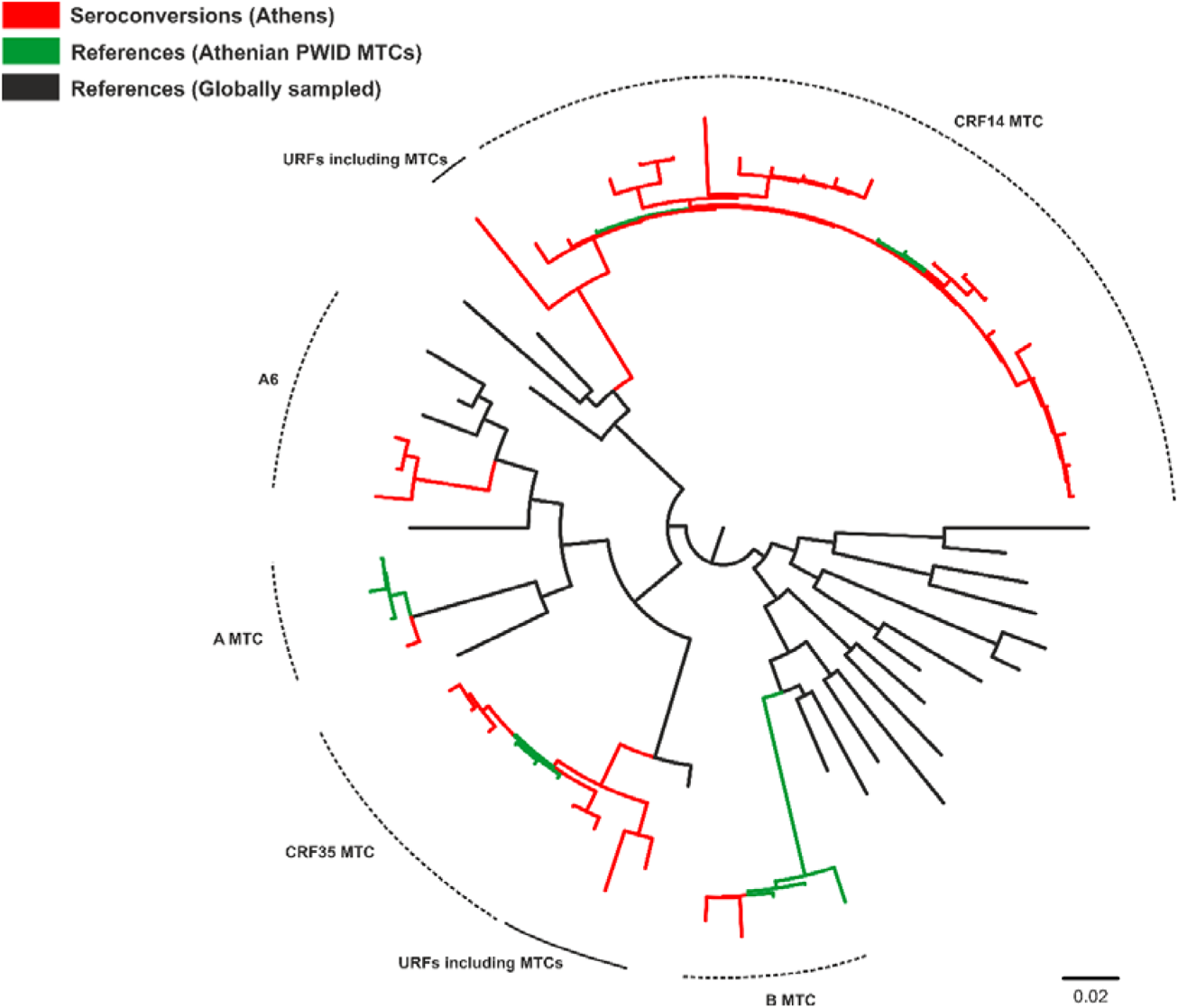
Unrooted phylogenetic tree of the HIV-1 sequences under study (seroconversions; red color) and reference sequences (black and green colors) inferred by using an approximately maximum-likelihood method as implemented in FastTree2 program. Representative sequences of pure HIV-1 subtypes, sub-subtypes and circulating recombinant forms collected from the Los Alamos HIV-1 sequence database (marked in black) and sequences from the four major molecular transmission clusters among people who inject drugs in Athens, Greece, (marked in green) were used as references. For the sake of clarity, a subset of the reference sequences was used. The names of subtypes, sub-subtypes, circulating recombinant forms, and unique recombinant forms are shown on the top of the corresponding clades. Abbreviations: CRF, circulating recombinant form; MTC, molecular transmission cluster; PWID, people who inject drugs; URF, unique recombinant form.

**Figure 2B.**
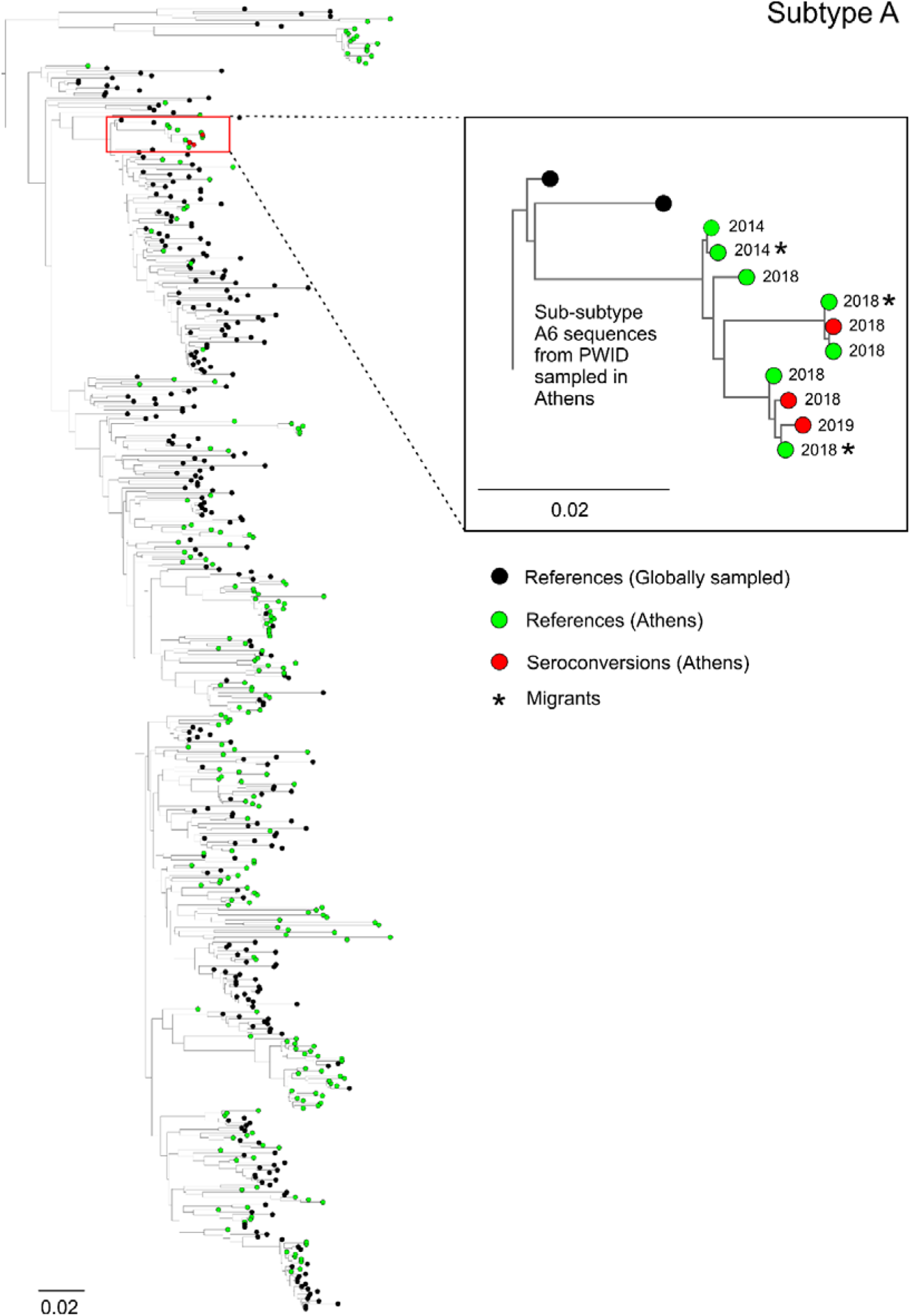
Unrooted phylogenetic tree of HIV-1 subtype A sequences from Athens, Greece, and other geographic regions around the world. For the sake of clarity, a subset of sequences was used. Red circles indicate seroconversions in Athens while green and black circles indicate sequences sampled in Athens and elsewhere, respectively. An enlarged view of the tree shows the sub-subtype A6 molecular transmission cluster consisted of 10 sequences from people who inject drugs sampled in Athens during 2014-2019. Asterisks indicate sequences from non-Greek people who inject drugs. The phylogenetic tree was inferred by using an approximately maximum-likelihood method as implemented in FastTree2 program.

## DISCUSSION

In the population of PWID in Athens, there was a deterioration of socioeconomic conditions in 2018-2020 compared to 2012-2013. Although heroin remained the main substance injected by most participants, there was a shift in the use of cocaine, a substance associated with increased risk of HIV infection and outbreaks (26–28). Encouragingly, there was a shift to less frequent injecting drug use. This is in agreement with data from people entering drug treatment programs in Greece, where users of opioids as well as of cocaine and other stimulants reported injection in 2019 at a lower frequency as compared to 2012 (29). Although in 2018-2020 there was increased access to OST programs, access to free syringes decreased, and HIV testing rates remained low (around half tested in the past 12 months).

HIV prevalence per round in 2018-2020 was approximately 11% and at similar levels as in 2012-2013 (4). However, in the sub-sample of PWID who had participated in both programs, HIV prevalence increased in 2018-2020 by approximately 55% as compared to 2012-2013 (from 14.2% to 22.0%). These findings are not contradictory. The first approach is based on cross-sectional data collected at multiple time points and the prevalence estimates reflect the situation in the target population in that time period. The prevalence cohort allows to track the same participants in the two time periods and to assess whether there is ongoing transmission. From an HIV surveillance point of view, these results provide evidence that repeated-cross sectional surveys cannot adequately capture transmission dynamics and that cohort studies with active enrollment are necessary as surveillance tools.

In a previous analysis of the 2011 outbreak, we estimated that HIV incidence decreased rapidly from 7.8/100 PYs in August-December 2012 to 1.7/100 PYs in August-December 2013 (4). In the current study, we found that HIV incidence remained at stable moderate levels, similar to those reached in the second half of 2013. Phylogenetic analysis showed that the majority of the viral sequences from seroconverters fell within the previously identified PWID molecular transmission clusters in Athens, or they were recombinants with viruses belonging to these clusters (17). Notably, besides the transmissions originating from the existing outbreak clusters, almost 9% of PWID seroconversions occurred within a newly expanding A6 cluster. The earliest sequences of this cluster were from 2014 and probably introduced from Eastern Europe, where this sub-subtype is endemic. The generation and expansion of a new transmission cluster indicates that the PWID epidemic was fuelled with new strains from former Soviet Union countries (30–32) and with existing contacts among active PWID in Athens.

The most important drug-related risk factor for HIV seroconversion was daily injection. This is consistent with the findings concerning the drivers of transmission in the 2011 outbreak in Athens where daily injection was the only drug-related risk factor identified (4). Younger age was independently associated with increased risk of HIV seroconversion, as in other settings (33–36). We also identified lower educational level – a correlate of lower socioeconomic status – to be an independent risk factor, as elsewhere (37–40). Socio-economic factors have been proposed to trigger the Athens outbreak in the first place; Greece entered a phase of economic recession in 2008, and this resulted in increased socioeconomic disparities, unemployment, and homelessness among PWID (5).

An important question is why HIV transmission did not decline further after 2013. In other settings with increased transmission among PWID, such as Haiphong and Bangkok, HIV incidence declined in the recent years to <1/100 PYs (41, 42). In Athens, as soon as the HIV outbreak was recognized in 2011, there was a scale-up of NSP and OST. In addition, ARISTOTLE program reached rapidly a large proportion of PWID (estimated population coverage: 88%), offered HIV testing, counselling, and linkage to antiretroviral treatment (4, 7). Data on the cascade of care among people living with HIV in 2013 in Greece confirm that a high proportion of HIV-infected PWID were diagnosed (87%); however, only 46% initiated antiretroviral treatment (43). Following 2013, NSP coverage in Athens was far from optimal with fluctuations over time; from 216 syringes/PWID per year in 2013, it declined to 109 in 2015 and remained low at 164 in 2018 (44). In addition, after the completion of ARISTOTLE in 2013, there was no systematic HIV testing of PWID. Our data further confirm that access to free syringes and HIV testing remained suboptimal in both periods. The low levels of testing and NSP coverage in a high HIV prevalence population that has experienced a recent outbreak is alarming. An increase in access to OST was observed in 2018-2020 as compared to 2012-2013, but the coverage was suboptimal (22.9% for all participants and 30.5% in the prevalence cohort). There is a core of PWID not accessing harm reduction programs who are most vulnerable. Ideally, this population should be protected from HIV infection through counselling and education, adequate syringe provision, access to HIV testing and linkage to antiretroviral treatment implemented through community-based programs specifically designed to address their needs.

Our analysis was based on data collected from PWID recruited though community-based programs using peer-driven chain referral. As a result, our samples consist of high-risk PWID, predominantly current injectors and not linked to OST programs, i.e. a key population at risk of contracting and transmitting HIV and with low access to prevention, care, and treatment services. The population coverage of the two programs was high. The official capture-recapture population size estimate (95% CI) for the number of people injecting drugs in the past 30 days was 3,069 (2,520 – 3,797) in 2012 and 1,487 (861 – 3,104) in 2018 (44, 45). In our programs, we recruited 2,689 and 1,224 PWID reporting injection in the past 30 days. Thus, the population coverage (95% CI) was 88% (71%-100%) and 82% (39%-100%) in ARISTOTLE 2012-2013 and ARISTOTLE HCV-HIV 2018-2020, respectively.

Our analysis has some limitations. First, concerning the estimation of HIV incidence, there was no regular follow-up for 46 seroconversions with a negative test result in 2012-2013 identified positive in 2018-2020. To deal with this problem, for 30 out of 46 of these cases who were already reported to the national HIV surveillance system with an earlier diagnosis date, we used that date as a putative first positive date. As the midpoint approach might lead to an artefactual clustering of seroconversion times in the middle of the analysed period (13), we used as sensitivity analysis the random-point method that has been proposed for low testing rates in cohorts (13) and the results were similar. Second, HIV incidence was assessed in a sample of seronegative PWID who had at least two participations. It could be argued that this sample comprised more vulnerable PWID in need of the monetary incentives and, thus, HIV incidence might be over-estimated. However, in another paper, we estimated HIV incidence during the Athens outbreak using this approach as well as by applying the Limiting Antigen Avidity assay to all HIV positive participants (i.e. including those with at least one visit to the program) and the results were similar (46).

In conclusion, our analysis revealed that, compared to 2012-2013, there was deterioration of socioeconomic conditions among PWID in Athens in 2018-2020, a shift in the use of cocaine, reduced access to NSP and stable low levels of HIV testing. On the positive side, there was a shift to less frequent injecting drug use and increased access to OST programs. Although HIV prevalence remained overall stable in the two periods, the increase in HIV prevalence among participants tested in both periods, the identification of a new expanding phylogenetic cluster among seroconverters and the estimated HIV incidence reveal ongoing transmission with PWID of younger age, lower educational level, larger injection networks and daily injecting being most at risk. It should be noted that ARISTOTLE HCV-HIV was prematurely discontinued in February 2020 due to the COVID-19 pandemic. Preliminary data collection in Athens and sites with recent HIV outbreaks suggests that COVID-19 has severely impacted HIV prevention services for PWID, possibly resulting in an increased risk for HIV transmission among PWID (47). The ongoing

HIV transmission among PWID in Athens provides empirical evidence that the current level of prevention is inadequate to control the epidemic and results in the expansion of the pool of infected PWID. Re-evaluation of prevention and treatment programs is urgently needed.

## Data Availability

Data are not available for sharing.

## ACKNOWLEDGEMENTS

We would like to thank the participants of the two programs. We would also like to acknowledge the contribution of the staff of ARISTOTLE and ARISTOTLE HCV-HIV programs:

C. Bagos, M. Esmaili, H. Malekian, E. Karamanou, F. Leobilla, C. Mourtezou, E. Sidrou, M. Zigouritsas, M. Dimitropoulou, N. Kagelari, M. Michail, S. Papadopoulos, and A. Vlahos (*ARISTOTLE staff*)

M. Dragasaki, K. Kourousi, G. Antoniou, H. Malekian, P. Tzimis, N. Fitsialos, L. Polychronopoulou, E. Kokolesis, P. Axaopoulos, and K. Kontsantinou (*ARISTOTLE HCV-HIV staff)*

ARISTOTLE HCV-HIV was conducted under the auspices of a) the City of Athens and b) the Organization Against Drugs (OKANA). We would like to acknowledge the support of the following persons in the implementation of this program: the Mayor of the City of Athens (K. Bakoyannis), K. Kokkolis and A. Theocharis (Organisation Against Drugs) and A. Fotiou (Greek National Monitoring & Documentation Center for Drugs & Drug Addiction).

## PREPRINT STATEMENT

A preprint of this article has been posted at https://www.medrxiv.org/content/10.1101/2021.06.24.21258830v1.

## AUTHORS CONTRIBUTIONS

Sotirios Roussos: Data Curation; Formal Analysis; Methodology; Project Administration; Software; Visualization; Writing – Original Draft Preparation; Writing – Review & Editing

Dimitrios Paraskevis: Conceptualization; Formal Analysis; Methodology; Project Administration; Supervision; Resources; Visualization; Writing – Original Draft Preparation; Writing – Review & Editing

Mina Psichogiou: Investigation; Writing – Review & Editing

Evangelia Georgia Kostaki: Data Curation; Formal Analysis; Software; Visualization; Writing – Review & Editing

Eleni Flountzi: Data Curation; Formal Analysis; Writing – Review & Editing

Theodoros Angelopoulos, Savvas Chaikalis: Investigation; Writing – Review & Editing

Martha Papadopoulou: Data Curation; Investigation; Writing – Review & Editing

Ioanna D Pavlopoulou: Writing – Review & Editing

Meni Malliori: Resources; Writing – Review & Editing

Eleni Hatzitheodorou, Magdalini Pylli, Chrissa Tsiara, Dimitra Paraskeva: Investigation; Writing – Review & Editing

Apostolos Beloukas: Formal Analysis; Resources; Writing – Review & Editing

George Kalamitsis: Funding Acquisition; Resources; Writing – Review & Editing

Angelos Hatzakis: Conceptualization; Funding Acquisition; Methodology; Resources; Project Administration; Supervision; Writing – Original Draft Preparation; Writing – Review & Editing

Vana Sypsa: Conceptualization; Formal Analysis; Funding Acquisition; Methodology; Project Administration; Supervision; Writing – Original Draft Preparation; Writing – Review & Editing

