## Supplementary appendix for "Ongoing HIV transmission following a large outbreak among people who inject drugs in Athens, Greece (2014-2020)"

**Comparison of PWID who participated in both programs vs. those who participated only in 2012-2013.**

To assess differential loss to follow-up, we compared the characteristics of PWID who participated in both programs versus those who participated only in ARISTOTLE 2012-2013. Compared to the total sample of participants of ARISTOTLE 2012-2013, repeat participants were younger (mean (SD): 35.0 (7.5) vs. 36.0 (8.5), p=0.004), more frequently females (18.7% vs. 14.6%, p=0.010), of Greek origin (94.9% vs. 81.4%, p<0.001), with stable accommodation (83.8% vs. 75.1%, p<0.001), with history of imprisonment (21.9% vs. 17.7%, p=0.013), reported more often sharing syringes in the past 12 months (40.8% vs. 35.9%, p=0.018) and having been tested for HIV within the past year (50.9% vs. 44.6%, p=0.011) (Supplementary Table 2).

**Supplementary Table 1.** Trends in socio-economic and network characteristics, drug use behaviour and access to HIV prevention services in people who inject drugs based on ARISTOTLE (2012-2013) (N=3,320) and ARISTOTLE HCV-HIV (2018-2020) programs (N=1,635) (as assessed in their first visit to each program).

|  | **2012-2013**  **(N=3,320)** | **2018-2020**  **(N=1,635)** | **p-value** |
| --- | --- | --- | --- |
| **A. Socio-economic and network characteristics** |  |  |  |
| Homeless now, % (95% CI) | 23.1 (21.7-24.6) | 26.9 (24.7-29.1) | 0.004^1^ |
| Unemployment, % (95% CI) | 79.1 (77.7-80.5) | 91.0 (89.5-92.3) | <0.001^1^ |
| Country of origin: Greece, % (95% CI) | 83.6 (82.3-84.9) | 84.4 (82.6-86.2) | 0.464^1^ |
| Without health insurance, % (95% CI) | 64.1 (62.4-65.7) | 79.8 (77.7-81.7) | <0.001^1^ |
| History of imprisonment (past 12 months), % (95% CI) | 18.5 (17.2-19.9) | 13.3 (11.7-15.0) | <0.001^1^ |
| Size of participant’s injection network, median (25th, 75th) | 30 (10-50) | 20 (10-50) | <0.001^2^ |
| **B. Injecting drug use behaviour** |  |  |  |
| Main substance of use (past 12 months), % (95% CI) |  |  |  |
| Heroin/Thai | 81.9 (80.6-83.2) | 59.4 (56.8-61.8) | <0.001^1^ |
| Cocaine | 14.0 (12.8-15.2) | 23.7 (21.6-25.9) | <0.001^1^ |
| Speedball | 2.8 (2.3-3.4) | 15.5 (13.7-17.4) | <0.001^1^ |
| Other | 1.3 (0.9-1.7) | 1.4 (0.9-2.2) | 0.662^1^ |
| Injecting drug use (past 30 days), % (95% CI) | 81.2 (79.9-82.6) | 74.9 (72.7-76.9) | <0.001^1^ |
| Daily injecting drug use (past 12 months), % (95% CI) | 38.0 (36.4-39.7) | 28.9 (26.7-31.2) | <0.001^1^ |
| Receptive syringe-sharing about half the time or more (past 12 months), % (95% CI) | 7.6 (6.7-8.6) | 6.4 (5.3-7.7) | 0.129^1^ |
| Use drugs divided with a syringe that someone else had already injected with about half the time or more (past 12 months), % (95% CI) | 8.0 (7.1-9.0) | 7.4 (6.2-8.8) | 0.467^1^ |
| **C. Access to testing, drug treatment and prevention** |  |  |  |
| Currently in opioid substitution treatment, % (95% CI) | 12.5 (11.4-13.7) | 22.9 (20.9-25.0) | <0.001^1^ |
| Received free syringes (past 12 months), % (95% CI) | 50.6 (48.9-52.3) | 41.2 (38.8-43.7) | <0.001^1^ |
| Tested for HIV (past 12 months), % (95% CI) | 45.9 (44.2-47.6) | 52.2 (49.7-54.8) | <0.001^1^ |

^1^Chi-squared test, ^2^Mann–Whitney U test - Abbreviation: CI, confidence interval.

**Supplementary Table 2.** Comparison of characteristics of PWID participating in both programs (N=681) versus those participating only in ARISTOTLE 2012-2013 (N=2,639) to assess differential loss to follow-up (characteristics were assessed in their first visit in ARISTOTLE 2012-2013).

|  | **Both programs**  **(N=681)** | **Only in ARISTOTLE**  **2012-2013**  **(N=2,639)** | **p-value** |
| --- | --- | --- | --- |
| **A. Socio-demographic and network characteristics** |  |  |  |
| **Age (years)**, mean [95% CI] (SD) | 35.0 [35.7-36.4] (7.5) | 36.0 [34.4-35.6] (8.5) | 0.004^1^ |
| **Gender**, n [%; (95% CI)] |  |  | 0.010^2^ |
| Male | 554 [81.4; (78.2-84.2)] | 2253 [85.4; (84.0-86.7)] |  |
| Female | 127 [18.6; (15.8-21.8)] | 386 [14.6; (13.3-16.0)] |  |
| **Country of origin**, n [%; (95% CI)] |  |  | <0.001^2^ |
| Greece | 646 [94.9; (92.9-96.4)] | 2147 [81.4; (79.8-82.8)] |  |
| Other | 35 [5.1; (3.6-7.1)] | 492 [18.6; (17.2-20.2)] |  |
| **Highest completed level of education**, n [%; (95% CI)] |  |  | 0.342^2^ |
| Middle/secondary school or below | 402 [59.7; (55.9-63.5)] | 1509 [57.7; (55.8-59.6)] |  |
| High school or higher level | 271 [40.3; (36.5-44.1)] | 1106 [42.3; (40.4-44.2)] |  |
| **Homeless now**, n [%; (95% CI)] |  |  | <0.001^2^ |
| No | 570 [83.8; (80.8-86.5)] | 1977 [75.1; (73.4-76.8)] |  |
| Yes | 110 [16.2; (13.5-19.2)] | 655 [24.9; (23.2-26.6)] |  |
| **Without health insurance**, n [%; (95% CI)] |  |  | 0.153^2^ |
| No | 259 [38.3; (34.6-42.0)] | 927 [35.3; (33.5-37.2)] |  |
| Yes | 418 [61.7; (58.0-65.4)] | 1699 [64.7; (62.8-66.5)] |  |
| **History of imprisonment (past 12 months)**, n [%; (95% CI)] |  |  | 0.013^2^ |
| No | 529 [78.1; (74.8-81.2)] | 2164 [82.3; (80.8-83.8)] |  |
| Yes | 148 [21.9; (18.8-25.2)] | 465 [17.7; (16.2-19.2)] |  |
| **Size of participant’s injection network**, median (25th, 75th) | 20 (10, 50) | 30 (10, 50) | 0.131^3^ |
| **B. Injecting drug use behaviour** |  |  |  |
| **Main substance of use (past 12 months)**, n [%; (95% CI)] |  |  | 0.028^2^ |
| Heroin/Thai | 549 [80.7; (77.6-83.6)] | 2151 [82.2; (80.7-83.7)] |  |
| Cocaine | 113 [16.6; (13.9-19.6)] | 349 [13.3; (12.1-14.7)] |  |
| Speedball | 14 [2.1; (1.1-3.4)] | 78 [3.0; (2.4-3.7)] |  |
| Other | 4 [0.6; (0.2-1.5)] | 38 [1.5; (1.0-2.0)] |  |
| **Injecting drug use in the past 30 days**, n [%; (95% CI)] |  |  | 0.724^2^ |
| No | 124 [18.3; (15.4-21.4)] | 497 [18.9; (17.4-20.4)] |  |
| Yes | 554 [81.7; (78.6-84.6)] | 2135 [81.1; (79.6-82.6)] |  |
| **Duration of injecting drug use (years)**, median (25th, 75th) | 13 (8, 18) | 12 (6, 19) | 0.544^3^ |
| **Daily injecting drug use (past 12 months)**, n [%; (95% CI)] |  |  | 0.264^2^ |
| No | 434 [63.8; (60.1-67.4)] | 1619 [61.5; (59.6-63.4)] |  |
| Yes | 246 [36.2; (32.6-39.9)] | 1014 [38.5; (36.6-40.4)] |  |
| **Receptive syringe-sharing Sharing (past 12 months**, n [%; (95% CI)] |  |  | 0.018^2^ |
| Never or rarely | 402 [59.2; (55.4-62.9)] | 1682 [64.1; (62.2-65.9)] |  |
| About half the time or more | 277 [40.8; (37.1-44.6)] | 942 [35.9; (34.1-37.8)] |  |
| **Use drugs divided with a syringe that someone else had already injected with (past 12 months)**, n [%; (95% CI)] |  |  | 0.348^2^ |
| Never or rarely | 615 [91.1; (88.7-93.1)] | 2415 [92.2; (91.1-93.2)] |  |
| About half the time or more | 60 [8.9; (6.9-11.3)] | 204 [7.8; (6.8-8.9)] |  |
| **C. Access to testing, drug treatment and prevention** |  |  |  |
| **Currently in opioid substitution treatment**, n [%; (95% CI)] |  |  | 0.239^2^ |
| No | 579 [86.2; (83.3-88.7)] | 2284 [87.8; (86.5-89.1)] |  |
| Yes | 93 [13.8; (11.3-16.7)] | 316 [12.2; (10.9-13.5)] |  |
| **Received syringes through prevention activities (past 12 months)**, n [%; (95% CI)] |  |  | 0.470^2^ |
| No | 327 [48.2; (44.3-52.0)] | 1308 [49.7; (47.8-51.6)] |  |
| Yes | 352 [51.8; (48.0-55.7)] | 1323 [50.3; (48.4-52.2)] |  |
| **Tested for HIV**, n [%; (95% CI)] |  |  | 0.011^2^ |
| Within past year | 344 [50.9; (47.0-54.7)] | 1163 [44.6; (42.7-46.5)] |  |
| More than 1 year ago | 124 [18.3; (15.5-21.5)] | 511 [19.6; (18.1-21.2)] |  |
| Never tested | 208 [30.8; (27.3-34.4)] | 933 [35.8; (33.9-37.7)] |  |

^1^t-test, ^2^Chi-squared test, ^3^Mann–Whitney U test - Abbreviation: CI, confidence interval.

**Supplementary Table 3.** Trends in HIV prevalence per RDS round in people who inject drugs in 2012-2013 and 2018-2020 programs in Athens, Greece (all participants included). The estimates for the period 2012-2013 were obtained from (1).

|  | **2012-2013** | | | | | **2018-2020** | |
| --- | --- | --- | --- | --- | --- | --- | --- |
| **Round** | **A** | **B** | **C** | **D** | **E** | **A** | **B** |
| HIV(+)/N | 275/1404 | 237/1438 | 247/1429 | 218/1406 | 203/1401 | 218/1361 | 90/576 |
| HIV prevalence^1^ (%) | 14.2 | 16.1 | 16.2 | 13.5 | 12.0 | 10.7 | 11.3 |
| (95% CI) | (10.3, 18.0) | (12.0, 20.1) | (11.9, 20.4) | (9.5, 17.5) | (8.6, 15.5) | (8.2, 14.4) | (8.0, 16.7) |

^1^ RDS-weighted estimates

1.	Sypsa V., Psichogiou M., Paraskevis D., Nikolopoulos G., Tsiara C., Paraskeva D. et al. Rapid Decline in HIV Incidence Among Persons Who Inject Drugs During a Fast-Track Combination Prevention Program After an HIV Outbreak in Athens, J Infect Dis 2017: 215: 1496-1505.

**Supplementary Figure 1.** Adjusted −ln{−ln(survival)} curves against time for each category of the identified risk factors for HIV seroconversion

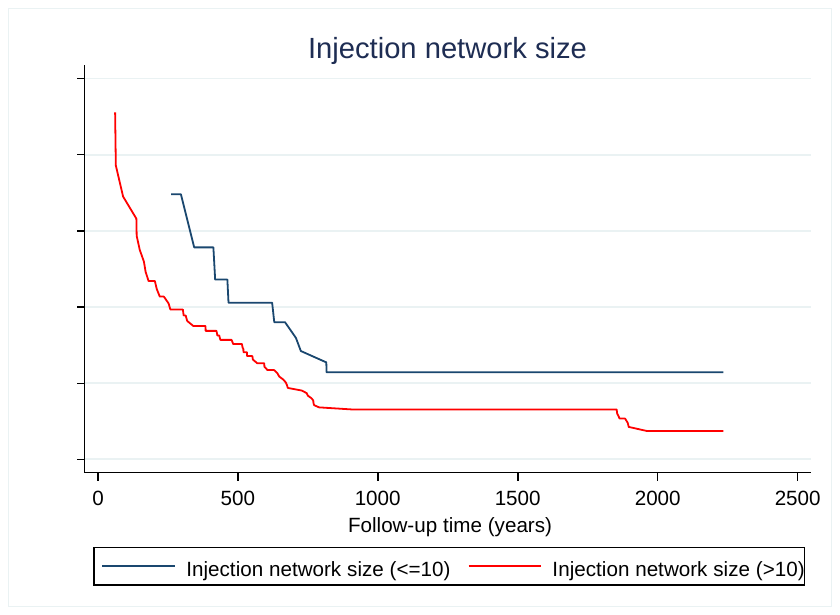

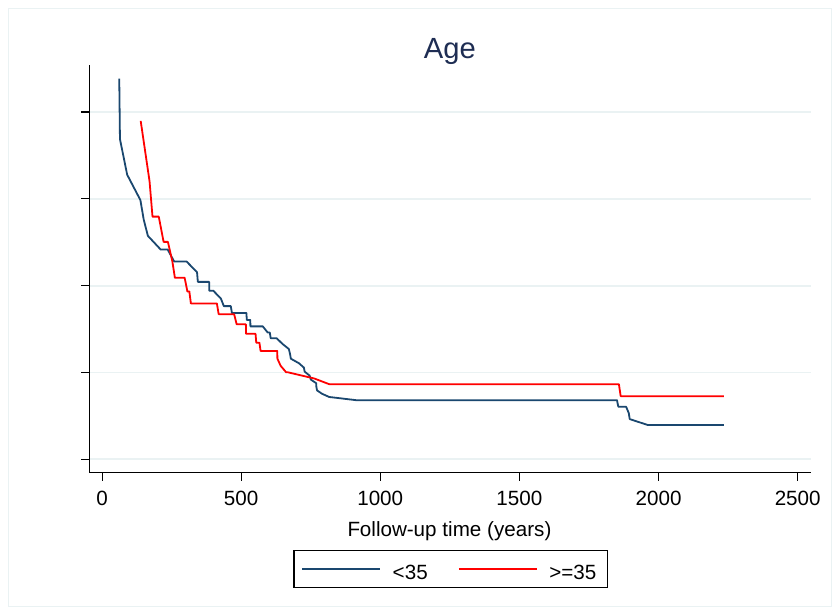

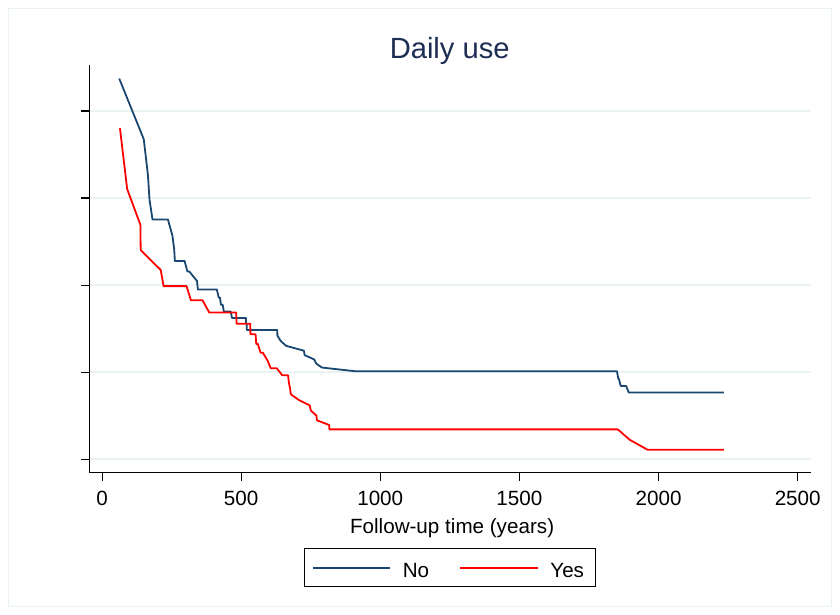

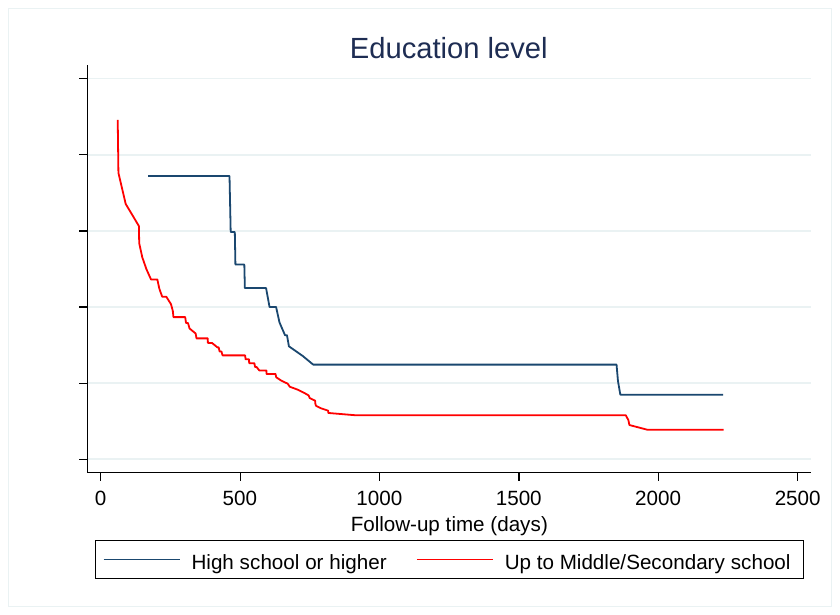
